# Transformers in Skin Lesion Classification and Diagnosis: A Systematic Review

**DOI:** 10.1101/2024.09.19.24314004

**Authors:** Abdulmateen Adebiyi, Nader Abdalnabi, Eduardo J. Simoes, Mirna Becevic, Emily Hoffman Smith, Praveen Rao

**Affiliations:** Department of Electrical Engineering and Computer Science, University of Missouri,United States; MU Institute for Data Science and Informatics, University of Missouri, United States; Department of Biomedical Informatics, Biostatistics and Medical Epidemiology, University of Missouri, United States; Department of Dermatology. University of Missouri, United States; Department of Dermatology, Saint Louis University, United States

**Keywords:** Transformer, Vision Transformer, Skin Cancer, Primary Care Physicians

## Abstract

**Objectives:** This systematic review aims to provide a comprehensive overview of the current state of research on the application of transformers in skin lesion classification.

**Materials and Methods:** Over the period 2017-2023, this systematic review investigated the application of transformer-based models in skin lesion classification, focusing on 57 articles retrieved from prominent databases which are PubMed, Scopus, and Medline. The inclusion criteria encompass studies centering on transformer-based models for skin lesion classification, utilization of diverse datasets (dermoscopic images, clinical images, or histopathological images), publication in peer-reviewed journals or conferences, and availability in English. Conversely, exclusion criteria filter out studies not directly related to skin lesion classification, research applying algorithms other than transformer-based models, non-academic articles lacking empirical data, papers without full-text access, and those not in English.

**Results:** Our findings underscore the adaptability of transformers to diverse skin lesion datasets, the utilization of pre-trained models, and the integration of various mechanisms to enhance feature extraction.

**Conclusion:** Our Systematic review showed the implementation and the application of the Transformer models for skin lesion classification. Our study showed the research areas and future research ideas for the use of transformers on skin lesion classification.

## Introduction

Skin cancer is the most common type of cancer in the United States^1^. Melanoma, a type of skin cancer, accounts for most skin cancer deaths^1^. About 100,640 new cases of melanoma will be diagnosed in 2024^1^. Around 59,170 of these cases are in men and 41,470 in women^1^. Specifically, the unregulated growth of abnormal skin cells is identified as skin cancer. DNA damage in skin cells from exposure to Ultraviolet (UV) radiation from the sun or tanning beds leads to mutations or genetic abnormalities. These mutations prompt the rapid reproduction of skin cells^2^.

The initial stage in diagnosing a potentially malignant skin lesion involves a visual inspection by a trained clinician^3^. Achieving an accurate diagnosis is crucial due to the similarities among other lesion types, such as seborrheic keratosis, pigmented spindle cell nevus, among others.^2^ Notably, the Computer-Aided System (CAD) demonstrates a diagnostic accuracy closely comparable to that of an experienced dermatologist^4^. In the absence of technological support, dermatologists with over 10 years of clinical experience can diagnose melanoma with an accuracy rate of around 65% to 80%^5,6^. In cases requiring additional verification, dermatoscopic images are captured using a high-resolution camera to supplement the visual examination^7^. This process involves controlled lighting and the use of a filter during image capture to diminish skin reflections^8^.Integration of this technical assistance has yielded a significant improvement in skin lesion diagnosis. Ultimately, the synergy between visual examinations and dermatoscopy images have resulted in an improved accuracy ranging from 75% to 84% for melanoma detection^9^.

The emergence of the transformer models has sparked a lot of interest in the vision community because of their great performance on natural language tasks^10,12^. Unlike previous methods like gated Neural Networks (NNs), Recurrent Neural Networks (RNNs), and Long-Short Term Memory (LSTM) networks, transformers enables global dependency modeling and parallelization^11,12^.

Many of the available reviews on the detection of skin lesion are focused on Convolution Neural Networks^2,13,14^. With the recent introduction of the transformers model, researchers have started implementing transformer models for skin lesion detection. Papa et al. systematic review on efficient vision transformers focused on the efficient methodologies for optimal estimation performance^15^. They analyzed efficient strategies like compact architecture, pruning methods, knowledge distillation and quantization. Our work is different from existing reviews as we surveyed the transformer models implemented in different manuscripts and its applications for skin lesion detection.

*Our objectives in this systematic review are:*

- To analyze current research on the use of transformers in skin lesion classification.
- To compare the performance of the different transformer models with convolution neural network architecture in skin lesion classification.
- To discuss the research gaps, future research ideas, under-researched areas and opportunities regarding the use of transformers on skin lesion classification.

## Methods

### Search process

We utilized three databases—PubMed, Scopus, and Medline—to retrieve pertinent research articles. The search was conducted in July 2024, and all obtained search results were considered. We examined the reference lists of the finalized articles to ensure any additional relevant studies are included. Our search string encompassed two major terms: “Skin Neoplasms” and “Transformers.” Variations of the term “Skin Neoplasms” included “Cancer of Skin” and “Skin Cancer,” while for “Transformers,” we employed alternate phrasings such as “Vision Transformers” and “Image Transformers.” The search strategy involved using diverse forms of each term and refining the search string based on both search outcomes and database specifications. PRISMA guidelines are followed for this scoping review. Figure 1 shows the flow of the PRISMA guidelines.

### Inclusion and exclusion criteria

Our inclusion criteria included articles that reported transformer-based approaches for skin lesion detection using dermascopy images, published in the English language, and in or after 2017. We included research studies that used transformers for lesion segmentation, lesion boundaries identification, lesion detection, and semantic information/features calculation from dermoscopy images. We excluded studies that used transformers for medical image data other than skin cancer applications.

### Study selection

In this study, we employed the PRISMA tool for the initial screening and selection of studies. Duplicates were removed, and the remaining studies were evaluated based on their titles and abstracts. The contents of the studies that met the inclusion and exclusion criteria were then assessed for eligibility.

### Data extraction

A data extraction sheet was prepared to retrieve all relevant information from the final included articles. This information includes the first author’s name, publication year, type of article, first author’s institution and location (country), data modality, availability of data (public or private, with access link), and architecture of the transformer model.

## Results

After applying the inclusion and exclusion criteria, we identified 20 papers focused on implementation of transformers architecture for skin lesion classification.

Summary Table in the supplementary materials shows the summary of the selected papers for review

### Dataset

Majority of the reviewed studies (n=17) used publicly available dataset. The HAM10000 dataset was the most widely used dataset in the systematic review. Nie et al implemented their hybrid model on the ISIC 2018 dataset^16,17^. They split the dataset into training, validation and testing set in the ratio 7:2:1. Aladhadh et al used the HAM1000 dataset^18^. They divided the dataset in to three parts for their experiment. The model was trained on 70% of the HAM10000 dataset, validated their model on 20% of the dataset and tested on 10% on the dataset.

Xin et al adopted two datasets for their study^19^. They used the HAM10000 dataset, and a custom dataset collected through dermoscopy. Desale et al also applied their vision transformer model on a publicly available dataset^20^; the International Skin Imaging Collaboration 2019 (ISIC 2019). They worked on a publicly available dataset^19^; the HAM10000 dataset. Different data augmentation like Horizontal_flip was used in this study, Vertical_flip,Random_crop,Random_rotation and color_jitter.

Abbas et al combined a set of four different publicly available datasets (Ph2, ISBI-2017, HAM10000 and ISIC) for their skin lesion classification.

Cirrincione et al used the publicly available ISIC 2017 in their studies too^21^. Their training dataset has 2000 lesions, the validation dataset has 150 images, and the testing set has 600 images. Yacob et al in their work applied their transformer model on a custom dataset that were collected at Sahlgrenska University Hospital, Gothenburg, Sweden from 2019 to 2020^22^, their dataset had a total of 1832 whole slide images (WSIs) from 479 Basal cell carcinoma (BCC). The training set is 1435 WSIs from 369 BCCs and testing set of 397 WSIs from 110 BCCs. Wang et al worked on two skin lesions segmentation datasets: The ISIC-2016&PH ^23^and the ISIC-2018. The ISIC-2016&PH2 contains samples from ISIC-2016 with 900 training samples and 379 datasets for validation. The PH2 dataset contains 200 lesions. The ISIC-2016 dataset was used for their training, and they validated their model on the PH2 dataset.

Guang et al. worked on the publicly available HAM10000 dataset and the Edinburgh DERMOFIT dataset^24^. In their experiment, 85% of the dataset was used for training and 15% of the dataset was used for testing. Saliva et al employed the Hyperspectral (HS) dataset in their work for the skin lesion classification. The HS dataset has 76 lesions from 61 subjects^25^. 46 of the lesions are malignant and 30 are benign.

Vachmanus et al evaluated their model on four publicly available datasets^26^. They are the PH2, SKINL2, PAD-UFES-20 and ISIC-20 datasets. Gulzar et al used the publicly available ISIC 2018 for their segmentation task^27^. The dataset has 2594 images for training and 1000 images for testing. Lungu-stan et al employed the ISIC 2019 dataset in their work. Figure 2 shows some of the sample images of the HAM10000 dataset.

The table below briefly describes the datasets that are in the different manuscripts in our systematic review.

**Table 1:** Brief Description on the datasets of the reviewed papers.

| Dataset | Year Released | Image Size | Number of Classes |
| --- | --- | --- | --- |
| ISIC-2016 | 2016 | 1279 | 2 (Malignant, Benign) |
| ISIC 2017 | 2017 | 2600 | 4 (Melanoma, nevus or seborrheic keratosis, seborrheic keratosis and melanoma or nevus) |
| HAM10000 (ISIC 2018) | 2018 | 10015 | 7 (AK, BCC, BKL, DF, NV, MEL, VASC) |
| ISIC 2019 | 2019 | 33569 | 8 (MEL, NV, BCC, AKIEC, BKL, DF, VASC, SCC) |
| ISIC 2020 | 2020 | 44108 | 2 (Malignant, Benign) |
| ISBI-2017 | 2017 | 2750 | 3 (NV, Mel, and SK) |
| Custom Data | Not Reported | 1016 | 3 (Basal Cell carcinoma, Squamous cell carcinoma and Malignant melanoma) |
| Ph2 | 2013 | 200 | 3 (nevus, atypical and melanomas) |
| Custom Dataset<br>(Sahlgrenska University<br>Hospital, Sweden) | Not Reported | 1831 | 2 (Tumor or no Tumor)<br>3 (No Tumor, low-risk, and high-risk tumor)<br>5 classes (no tumor, and 4 grades of BCC; low aggressive superficial, low aggressive nodular, medium aggressive and highly aggressive, in line with the Swedish classification system) |
| Dermofit dataset | Not Reported | 1300 | 10 (Actinic Keratosis, Basal Cell Carcinoma, Melanocytic Nevus, Seborrhoeic Keratosis, Squamous Cell Carcinoma, Intraepithelial Carcinoma, Pyogenic Granuloma, Haemangioma, Dermatofibroma, Malignant Melanoma) |
| PAD-UFES-20 | 2020 | 2298 | 6 (Basal Cell Carcinoma, Squamous Cell Carcinoma, Actinic Keratosis, Seborrheic Keratosis, Bowen's disease, Melanoma) |
| SKINL2 | 2019 | 814 | 8 (Melanoma, Melanocytic Nevus, Basal Cell Carcinoma, Seborrheic Keratosis, Hemangioma, Dermatofibroma, Psoriasis) |

Performance metrics Table in the supplementary materials shows the Performance metrics used in the reviewed papers.

### Transformers for Skin Lesion Classification

#### Transformer Architecture

Many of the papers we reviewed implemented models with transformer or transformer-like architecture^16,18,20^.Transformer architecture proposed by Vaswani et al^28^. as shown in figure 3 has the encoder-decoder structure. The encoder maps an input sequence which is then fed to the decoder. The decoder then generates the output sequence. They proposed a mechanism known as the Self Attention. Transformer has stacks of transformer blocks that maps sequences of input vectors (x1......xn) to sequences of output vectors (z1.... zn) of corresponding size. All these blocks are made by combining the simple linear layers, feedforward networks and self-attention layers. The transformer encodes the input sentence to sequence of vectors. The transfomer does the encoding by using the self-attention mechanism, that enables the model to learn the relationships between the words in the sentence^29^.

#### Vision Transformer

More specifically, some of the papers we reviewed implemented the Vision Transformer model. This is an image classification deep learning model that employs a transformer-like architecture for the classification task. The model was proposed in the research work entitled “An image is worth 16*16 words: Transformers for image recognition at scale”^30^. Vision transformers has been applied in many real-life applications like object detection, image classification etc. It has also been recently applied in many skin lesion classification tasks. Figure 4 shows the architecture for vision transformers.

Many of the papers we reviewed implemented the Vision Transformer model. Nie et al 2022 proposed a hybrid model that combines Convolution Neural Network (CNN) feature extraction with Vision transformer on the ISIC 2018 datasets^16^. Their Vision Transformer contains the embedding layer, the transformer encoder and the MLP head. They used ResNet50 for the feature extraction. The main difference between their model and the conventional Transformer model is that they used the traditional convolution neural network feature extraction with Vision Transformer. They performed their experiment using different loss function namely: Cross-Entropy Loss (CE), Weighted Cross-Entropy Loss (WCE) and Focal Loss (FL). They also compared the result of their Hybrid model (CNN+VIT) to ResNet50(CNN) model. They performed 6 different experiments using the loss function. The different experiments are using the ResNet50 model with the three Loss Function which they name (CNNce, CNNwce and CNNfl) and also using the Hybrid model (CNNViT) on the different loss functions which they name (CNNViTfc, CNNViTwce and CNNVITfl). Aladhadh et al proposed a Medical Vision Transformer (MVT) model for the Skin Cancer Classification^18^. The MVT architecture has the Embedding layer, Encoder Layer, and the Classification layer. Their model used an input data of the size 72 * 72. Their input image was converted into 9 patches. The MVT had 24 layers, hidden size D of 1024, MLP size of 4096 and a total parameter of 86 million.

Xin et al applied multiscale vision transformer (ViT) in their work^16^. A multiscale ViT improves the image feature embedding module. The major difference between their multi-scale vision transformer is that it mainly improves the image feature embedding module and applies the contrastive learning method to the skin cancer classification. The multi-scale sliding window generates the overlapping patch images. Their model has the multi-scale image sequentialization, the patch embedding and the contrastive learning.

Abbas et al proposed Assist Demo using a separable vision transformer to classify pigmented skin lesions^31^. Their model is based on the Squeezenet and depthwise separable convolution neural network models. They modified SqueezeNet design into Squeeze-light by using depthwise separable convolutions (SepConv). The SqueezeNet-Light architecture contains the SqueezeNet-Light base structure, fire module structure and SepConv Structure. They presented Assist-Dermo, a lightweight separable vision transformer model for classifying nine different classes of pigmented skin lesions (PSLs).

Cirrincione et al implemented a vision transformer model to classify the ISIC 2017 dataset^21^. Their proposed model is based on Vision Transformer which can be used for encoding images that are able to model long-range spatial relationships in the skin lesions. Their model has a Multi-Head Attention. The input is associated with key, query, and the value. Their result was fed to a SoftMax function and multiplied by the value vector. Their architecture encompasses patching, flattening, embedding and positional encoding, concatenation of the input sequence, layer normalization, multi-Head attention, layer normalization, Multilayer perceptron, classification head and the classification output.

Yacob et al proposed a weakly supervised approach for detecting and classifying basal cell carcinoma (BCC) using a graph-transformer on whole slide images (WSIs)^22^. They used features that were generated from self-supervised contrastive learning for their Graph Convolution Neural Network. They then fed the network to a Vision Transformer.

Wang et al used XBound-Former, a novel cross-scale boundary-aware transformer, for skin lesion segmentation from dermoscopy images^23^. X-Bound is a cross-scale attention mechanism for curbing the problems of ambiguous boundaries and size variation. The Cross-scale-aware transformer (XBound-Former) uses a pyramid vision transformer for extracting features of the skin lesions. The model addresses the variation and boundary problems of skin lesion segmentation by integrating boundary knowledge into a transformer-based network.

Vasu et al proposed an approach using Vision Transformers (ViT) to identify seborrheic keratosis skin disease, achieving 99% accuracy. The study compares ViT with VGG-19 and Inception-V3 CNN algorithms for classification^32^.

Yang et. al implemented a novel Vision Transformer model^21^. Their method has four bocks: The data augmentation and cancer-class rebalancing block, the image restructuring block, the transformer encoder block, and the classification block. They conducted four different experiments. They are ResNet50 with Soft Attention (ResNet50+SA), Inception ResNet with Soft Attention (Irv2+SA) and the two Vit for Skin Cancer detection (VitfSCD) models. The VitFSCD-base has 12 layers while the VITfSCD-large has 24 layers.

Salvia et al. proposed a novel hyperspectral image classification architecture utilizing Vision Transformers^25^. Their ViT model received N-D data (Hyperspectral dataset) that were transformed into a 1-D arrays by dividing the original image into patches of the same dimension. The method is validated on a real hyperspectral dataset containing 76 skin cancer images. Results demonstrate that Vision Transforms are suitable for this task, outperforming the state-of-the-art methods in terms of false negative rates and processing times. Additionally, the attention mechanism is evaluated for the first time on medical hyperspectral images.

Vachmanus et al. proposed DeepMetaForge, a deep-learning framework for skin cancer detection, which integrates metadata with visual features using a Vision Transformer backbone^26^. They used Bidirectional Encoder representation from Image Transformers (BEiT) backbone and the metadata forging mechanism. Their model contains the Image Encoder where they used the BEiT, the Metadata Encoder, which is based on a Convolution Neural Network, the Deep metadata fusion module that combines the image and metadata encoder and the classification layer.

Arshed at al proposed an approach using pre-trained Vision Transformers for multi-class skin cancer classification, addressing class imbalance within the dataset^33^. Their model preprocessed series of learned transformation of the input lesions. The input images were divided into 16*16 patches after resizing the images to 224 * 224 pixels.

Gulzar et al compares U-Net and attention-based methods for skin lesion image segmentation, aiming to assist in the diagnosis of skin lesions^27^. The hybrid TransUNet method is proposed and evaluated, achieving superior performance compared to benchmarking methods. The TransUNet architecture has been applied for medical image segmentation. TransUNet is a combination of the Transformer and U-Net architecture.

Lungu-Stan et al proposed a lightweight Vision Transformer for skin lesion classification, focusing on efficient inference and reduced memory usage. The model employs knowledge distillation and achieves competitive performance compared to larger models^34^. Khan et al proposed the SkinViT architecture^11^. They combined the idea of Yuan et al in VOLO^35^ and the VIT architecture^30^. Their model has the Outlooker block, transformer block and SkinViT Multilayer perceptron head block. The input images are transformed into an 8 by 8 patches which are then fed to the outlooker block. The tokens are then sampled using the patch embedding and then finally to the Multi-Layer Perceptron head. Overall, their proposed SkinViT method outperformed the State-of-the-art methods.

Wang et al proposed Melanoma BatchFormer Vision Transformer Model (MBViT) ^36^. The model combines the Vision Transformer model with the batchFormer module. They employed a dual-branch training strategy in their model training.

Reis al implemented a new deep learning model, a modified lightweight vision transformer (ViT) and a hybrid framework that combines an integrated deep learning model and an Ensemble learning model^37^. They named the model the multi head attention block depthwise separable convolution network (MABSCNET). The MABSCNET model was designed to combine the advantages of both the CNN and transformer model.

Dai et al introduced an efficient and lite convolution transformer model. Their model hierarchically learns the local and global representations through the attention mechanisms. Branch attention helps their model to extract features from different layers^34^.

Remya et al applied the Vision Transformer model for their skin lesion classification^35^. Each of their image was resized to 224 by 224 pixels and were set from 0 to 1 range. They applied EfficientNet-BO model as their feature extractor. A feedforward neural network was placed after many multi-head self-attention layers in their architecture.

Performance Table in the supplementary materials shows the performance metrics, the corresponding values in each paper, the models used in the papers and finally, the dataset

## Discussion

This systematic review analyzed different transformer techniques implemented and applied in skin lesion classification and diagnosis. The skin cancer detection process in reviewed manuscripts included the collection of the dataset, dataset pre-processing, the application of the transformer models and then the evaluation of the different transformer models using different performance metrics.

Most of the papers analyzed in this systematic literature review reported their best performance when Vision Transformer was applied on skin lesion dataset. They compared their performance with the state-of-the-art algorithms and other Convolution Neural Network as the benchmark. This shows that Transformer models specifically the Vision Transformer model generally outperforms the conventional convolution neural network models in the manuscripts we surveyed.

Nie et al Hybrid model outperformed the ResNet50 model with an accuracy of. 89.48%^13^. Specifically, they had the best performance when the Hybrid model (CNN+ViT) was combined with focal loss. Aladhadh et al Medical Vision Transformer (MVT) achieved an accuracy of 96.14%, precision of 96%, Recall/Sensitivity of 96.50% and F1-Measure of 97.00 ^15^. Xin et al proposed Multiscale VIT^16^. It achieved a precision of 0.942, an accuracy of 0.941 and an F1-score of 0.941 on their custom dataset and it achieved an AUC of 0.987, precision of 0.941 and an accuracy of 0.943 on the HAM10000 datasets. Abbas et al proposed the SqueezeNet-Light model^27^. It achieved a sensitivity of 94%, a specificity of 96%, an accuracy of 95.6%, a precision of 94.12% and an F1-Score of 95.2.

Cirrincione et al implemented a vision transformer model to classify the ISIC 2017 dataset^18^. Their model achieved an accuracy of 0.948, sensitivity of 0.928, specificity of 0.967 and AUROC of 0.948. Yacob et al proposed a weakly supervised approach for detecting and classifying basal cell carcinoma (BCC) using a graph-transformer on whole slide images (WSIs) ^19^. The approach achieved a high accuracy in both tumor detection and grading of BCCs, offering potential for increasing workflow efficiency in pathology laboratories. They achieved their best accuracy of 93.5% on the 2-class problem. Experimental results demonstrate the effectiveness of the proposed Wang et al used XBound-Former model, especially on boundary-wise metrics. Vasu et al proposed an approach using Vision Transformers (ViT) to identify seborrheic keratosis skin disease, achieving 99% accuracy^28^. The proposed method achieved high accuracy through class rebalancing, image preprocessing, and a transformer-based classification block. Yang et al. proposed VitfSCD-Large^21^. It achieved an overall accuracy of 94.1% on the HAM10000 dataset and VitfSCD-Base achieved an accuracy of 80.5% on the Dermofit dataset. Salvia et al. proposed a novel hyperspectral image classification that achieved a best accuracy of 93% on the malignant melanocytic dataset. Vachmanus et al. proposed DeepMetaForge, a deep-learning framework for skin cancer detection^23^. The method achieved high accuracy and outperformed CNN-based transfer learning models. It achieved an accuracy of 92.14%. Wang et al proposed Melanoma BatchFormer Vision Transformer Model (MBViT). Their proposed MBViT had the best performance with an accuracy of 97.73% on real images^36^. Reis et al implemented the multi head attention block depthwise separable convolution network (MABSCNET)^37^. The MABSCNET achieved the highest performance on the original dataset with an accuracy of 0.782. Dai et al introduced an efficient and lite convolution transformer model. Their model achieved an accuracy of 96.70%^38^.Remya et al applied the Vision Transformer model for their skin lesion classification. Their model achieved an accuracy of 99%, precision of 0.98, recall of 0.97 and an F1-score of 0.98^39^.

Dataset imbalance may lead to some biases in the evaluation of the skin cancer classification. Abbas et al pointed out that weighted cross entropy improves the performance of their models by correcting the class imbalance. Aladhadh et al. also handled the class imbalance problems in their preprocessing phase of the HAM10000 dataset^18^. They applied Brightness adjustment, Contrast Enhancement and Geometric Transformations in their Data Preprocessing phase^18^ They increased the numbers of AKIEC, BCC, DF and VASC to 1099 samples after preprocessing. They didn’t increase the numbers of the class BKL, NV and MEL. Nie et al 2022 handled the class imbalance problems in their dataset by using the loss functions: cross-entropy (CE), weighted-cross entropy (WCE), and Focal Loss function^16^. The Focal Loss strategy that Nie et al. implemented in their work was proposed by Lin et al^40^. Xin et al. employed label shuffling to handle the class imbalance in their experiments^16^.

Many of the studies we analyzed did not implement any explainability techniques like SHapley Additive exPlanations and Local Interpretable Model-agnostic Explanations. We strongly recommend that future research in the classification of skin lesions using transformer models to employ and prioritize model interpretability. Understanding the rationale behind model predictions is crucial for Primary Care Physicians and Nurse Practitioners, enabling them to make informed clinical decisions.

## Data Availability

All data produced in the present work are contained in the manuscript

## Acknowledgments

This project was funded by the Translational Research Informing Useful and Meaningful Precision Health (TRIUMPH) grant at the University of Missouri-Columbia.

